# Psychiatric polygenic risk scores: Child and adolescent psychiatrists’ knowledge, attitudes, and experiences

**DOI:** 10.1101/2021.10.08.21264763

**Authors:** Stacey Pereira, Katrina A. Muñoz, Brent J. Small, Takahiro Soda, Laura N. Torgerson, Clarissa E. Sanchez, Jehannine Austin, Eric A. Storch, Gabriel Lázaro-Muñoz

## Abstract

**Objective:** Psychiatric polygenic risk scores (PRS) have the potential to transform aspects of psychiatric care and prevention, but there are concerns about their implementation. We sought to assess child and adolescent psychiatrists’ (CAP) experiences, perspectives, and potential uses of psychiatric PRS.

**Methods:** A survey of 960 US-based practicing CAP.

**Results:** Most respondents (54%) believed psychiatric PRS are currently at least slightly useful and 87% believed they will be so in five years. Yet, 77% rated their knowledge of PRS as poor or very poor. Ten percent have had a patient/family bring PRS to them, and 25% would request PRS if a patient/caregiver asked. Respondents endorsed different actions in response to a hypothetical child with a top 5^th^ percentile psychiatric PRS but no diagnosis: 48% would increase prospective monitoring of symptoms, 42% would evaluate for current symptoms, and 4% would prescribe medications. Most respondents were concerned that high PRS results could lead to overtreatment and negatively impact patients’ emotional well-being.

**Conclusion:** Findings indicate emerging use of psychiatric PRS within child and adolescent psychiatry in the US. Thus, it is critical to examine the ethical and clinical challenges that PRS may generate and begin efforts to promote their informed and responsible use.

## Introduction

There have been recent advances in identifying genomic loci associated with a number of psychiatric conditions, including autism spectrum disorder, attention deficit hyperactivity disorder, schizophrenia, depression, and bipolar disorder.^1–7^ The identification of these genomic loci has made it increasingly possible to generate polygenic risk scores (PRS), which are weighted sums of risk across these genomic loci. PRS can then be used to stratify individuals by risk compared to the general population.^8^

Psychiatric PRS, most likely in combination with other predictors, have the potential to transform aspects of psychiatric care and prevention. The usual age of onset for many psychiatric conditions is during childhood, adolescence, or early adulthood,^9^ and as many as 20% of children and adolescents in the US have a diagnosable psychiatric disorder.^10^ Yet, diagnosis is often delayed, inaccurate, or does not take place, compounding impairment. As prediction tools, psychiatric PRS would be most useful before disorder onset. Initial studies suggest that psychiatric PRS-based estimates are promising. For example, an individual at the 99^th^ PRS percentile for schizophrenia or depression has a 6% or a 30% chance of developing these disorders, respectively.^11–13^ If utilized appropriately (e.g., ensuring any delivery of genetic test results is provided in the context of evidence-based psychiatric genetic counseling^14^), identifying those at increased risk could allow for more refined resource allocation toward those with the highest risk and potentially decrease the duration of untreated disorders, which is associated with improved clinical outcomes.^15,16^

Despite the potential for PRS to improve outcomes for those at risk, concerns about potential harms of psychiatric genetic testing in children have also been raised.^17^ Positive test results could lead to stigmatization, resulting in disruptions in familial or social relationships.^18^ For example, parents could potentially lower the degree to which they support their child’s educational, employment, or social aspirations.^19^ Stigmatization may also influence the formation of a child’s developing identity^20^ or minimize a child’s expectations for their future.^19^ Finally, the anxiety positive results may trigger could lead to the development of other psychiatric issues.^21–23^

Thus, critical clinical, ethical, and policy questions remain. For example, which children and adolescents, if any, should be tested and when, if at all, should children and adolescents be tested? In addition, the utility of PRS and how PRS should be used continue to be debated.^11,24–27^ Given the position of child and adolescent psychiatrists (CAP) as key stakeholders in the implementation of psychiatric PRS, it is critical to understand their perspectives toward use of PRS for psychiatric conditions in this population. Here we report CAP knowledge and experience with PRS, their perceived utility of PRS compared to other psychiatric genetic testing, their concerns about how psychiatric PRS could be used in child and adolescent psychiatry, how they anticipate they would respond to PRS results in their clinical practice, and their perception of the appropriateness of using psychiatric PRS for screening purposes. This study helps inform the clinical, ethical, and policy challenges raised by the use of PRS for psychiatric conditions.

## Methods

### Sampling

Survey participants were recruited from publicly available listservs, professional organizations, national conferences, and professional meetings. Web searches were conducted to identify other publicly available contact information for child and adolescent psychiatrists including social media, journal publications, and academic department websites. The survey was electronically distributed with two reminder emails over a four-week period in June 2020. Participants were compensated with a $10 gift card of their choice. The study was approved by the Baylor College of Medicine Institutional Review Board.

### Survey

A survey was developed to assess CAP current practice, knowledge, and perceptions towards genetic testing and was divided into three sections including general genetic testing, pharmacogenomics, and PRS. The 47-question survey (see supplementary materials) was developed based on current literature with input from an expert panel consisting of child and adolescent psychiatrists, psychologists, genetic counselors, bioethicists, lawyers, and an anthropologist using a modified Delphi method.^28^ Questions specific to PRS were used to learn about CAP knowledge, experience, current and potential utility, concerns, and appropriateness of PRS screening), the results of which we present here (see Soda et al.^29^ for results on knowledge and perceptions of utility of genetic testing in the evaluation of autism spectrum disorder). If participants selected “I have never heard of PRS” as the response to the initial self-reported knowledge of PRS question, display logic was used to skip participants to the next section without asking additional PRS questions. The entire survey took approximately 15-20 minutes to complete and was administered in English using Qualtrics.

#### Self-rated Knowledge of PRS

Respondents rated their knowledge of PRS by selecting one of the following response options: I have never heard of PRS, Very Poor, Poor, Good, or Very Good.

#### Experience

Respondents reported their experience with PRS in their clinical practice in four yes/no questions.

#### PRS Result Graph Interpretation

Respondents were presented with an example PRS result graph from impute.me^30,31^ and answered five questions about what the graph depicted for the individual’s risk of developing the condition.^32^ Response options included Agree, Disagree, and Unsure.

#### Potential Use of PRS

Respondents selected all applicable items among a list of 11 options about what would prompt them to request or generate a patient’s psychiatric PRS. They then indicated what they would do if a child or adolescent with no current psychiatric diagnosis had a high PRS for a psychiatric disorder by selecting all that applied among 13 items. Throughout the survey, “high PRS” was defined as “top 5%.” *Concerns*: Respondents indicated their level of concern about five potential negative outcomes of having a high (top 5%) psychiatric PRS result on a four-point scale with the descriptors: Not at All Concerned, Slightly Concerned, Somewhat Concerned, Very Concerned.

#### Appropriateness of Screening

Respondents indicated the degree of appropriateness to use PRS to screen in five groups and in two contexts on a four-point scale with the following descriptors: Very Inappropriate, Inappropriate, Appropriate, Very Appropriate. *Perceived Utility*: Respondents rated how useful they thought PRS are in child and adolescent psychiatry now and five years from now on a four-point scale with the descriptors: Not at all Useful, Slightly Useful, Moderately Useful, Very Useful.

### Analysis

Categorical variables were analyzed using chi-square and logistic regression. When cell sizes were small, Fisher’s Exact Test was used to compare the categorical groups. For comparisons of the current or future utility of PRS, McNemar change tests were computed.

## Results

### Participant Characteristics

The survey was electronically distributed to 5,677 participants via Qualtrics. Of the 1,180 (20.8%) who agreed to participate, 962 CAP completed the entire survey for an overall 16.9% completion rate, reflecting approximately 11.6% of CAP in current practice in the US.^29^ We then excluded two CAP who did not answer the self-rated knowledge question that determined whether they received the PRS section of the survey, for a final total of 960 respondents reported here (Figure 1).

**Fig 1.**
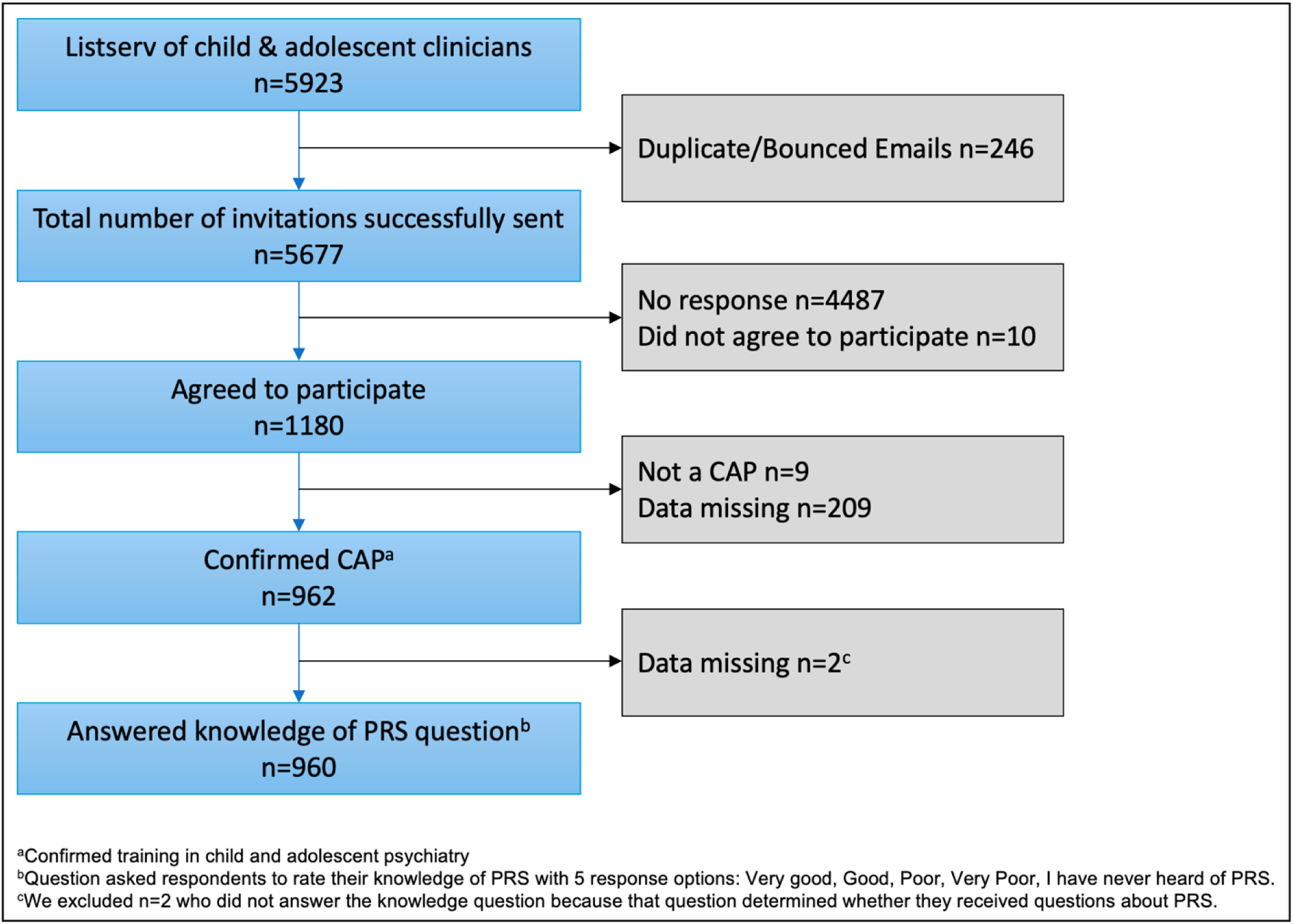
Study Flow

Participant characteristics for all 960 CAP are shown in Table 1. Among the CAP, 23.3% (n=223) indicated they had never heard of PRS. There were no differences in awareness of PRS as a function of gender, ethnicity, number of years of clinical practice, percentage of patients with ASD or IDD, or average age of patients. Lack of awareness of PRS was, however, associated with practice setting; 84.5% of CAP who reported that one of their practice settings was at a university medical center (UMC) were aware of PRS (OR=1.89, 95% CI=1.25 – 2.84, p=.003), as compared to persons who were not at a UMC (74.4%). Fewer CAP in private practice settings were aware of PRS (70.1%), as compared to those who did not list an affiliation with a private practice (80.5%; OR=.57, 95% CI=.42 – .77, p<.001). CAP who reported awareness of PRS (n=735) were then asked additional questions regarding knowledge, experience, perceived utility of, concerns about, and potential uses of this information in clinical practice and screening.

**Table 1.** CAPG Participant Characteristics

| **Participant Characteristics** | **Total Sample** | **Heard of PRS** | **Not Heard of PRS** | **Statistical Comparison** |
| --- | --- | --- | --- | --- |
| **Gender (n=956)** |  |  |  |  |
| Female | 476 (50) | 350 (74) | 125 (56) | $\chi^2 (4) = 6.23, p = .183$ |
| Male | 454 (47) | 361 (80) | 92 (41) |  |
| Prefer Not to Say | 26 (3) | 20 (77) | 6 (3) |  |
| Other | 1 (<1) | 1 (<1) | 0 (0) |  |
| Trans Male | 1 (<1) | 1 (<1) | 0 (0) |  |
| **Ethnicity (n=955)<sup>a</sup>** |  |  |  |  |
| White/ European American | 665 (70) | 506 (69) | 159 (71) | $\chi^2 (9) = 7.59, p = .576$ |
| Asian | 95 (10) | 75 (10) | 20 (9) |  |
| Prefer Not to Say | 54 (5) | 46 (6) | 8 (3) |  |
| Mixed Ethnicity/Race | 33 (4) | 25 (3) | 8 (3) |  |
| Black/African American | 37 (4) | 25 (3) | 12 (5) |  |
| Hispanic/Latinx | 36 (4) | 25 (3) | 11 (5) |  |
| Other | 17 (2) | 14 (2) | 3 (1) |  |
| Middle Eastern / Mediterranean | 13 (1) | 11 (1) | 2 (<1) |  |
| American Indian/Native | 4 (<1) | 4 (<1) | 0 (0) |  |
| American<br>Native Hawaiian/Pacific<br>Islander | 1 (<1) | 1 (<1) | 0 (0) |  |
| **Number Years in Clinical**<br>**Practice as CAP (n=960)** | | | | $\chi^2 (5) = 7.27, p = .201$ |
| 16+ years: | 532 (55) | 420 (57) | 112 (50) |  |
| 6-10 years: | 179 (18) | 136 (18) | 43 (19) |  |
| 11-15 years: | 167 (17) | 116 (16) | 51 (23) |  |
| 1-5 years: | 61 (6) | 47 (6) | 14 (6) |  |
| CAP Fellow: | 19 (2) | 15 (2) | 4 (2) |  |
| Resident: | 2 (<1) | 1 (<1) | 1 (<1) |  |
| **% Patients with ASD or**<br>**IDD (n=835)** | | | | $\chi^2 (4) = 5.42, p = .248$ |
| None or N/A | 33 (4) | 25 (4) | 8 (4) |  |
| 1-25% | 655 (79) | 486 (77) | 169 (83) |  |
| 26-50% | 101 (12) | 81 (13) | 20 (10) |  |
| 51-75% | 10 (<1) | 8 (1) | 2 (1) |  |
| 75-100% | 36 (4) | 32 (5) | 4 (2) |  |
| **Average % of Patients**<br>**(n=960)** | | | | $F(1, 957) = 0.00, p = .962$<br>$F(1, 957) = 1.46, p = .227$<br>$F(1, 957) = 2.42, p = .119$ |
| Under the Age of 12 | 28% | 28% | 28% |  |
| Ages 12-17 | 38% | 38% | 39% |  |
| Ages 18 and Up | 25% | 24% | 27% |  |
| **Main Practice Setting<sup>a</sup>**<br>**(n=955)** | | | | $\chi^2 (1) = 11.42, p < .001$<br>$\chi^2 (1) = 4.57, p = .033$<br>$\chi^2 (1) = 1.19, p = .275$<br>$\chi^2 (1) = 10.65, p = .001$<br>$\chi^2 (1) = 0.09, p = .763$<br>$\chi^2 (1) = 0.16, p = .694$<br>$\chi^2 (1) = 0.47, p = .492$<br>$\chi^2 (1) = 0.10, p = .752$<br>$\chi^2 (1) = 1.87, p = .171$<br>$\chi^2 (1) = 0.81, p = .368$ |
| Private Practice | 381 | 270 (71) | 111 (29) |  |
| Clinic | 267 | 217 (81) | 50 (19) |  |
| Hospital | 132 | 106 (80) | 26 (20) |  |
| University Medical Center | 227 | 192 (85) | 35 (15) |  |
| Community Agency | 114 | 86 (75) | 28 (25) |  |
| Psychiatric Hospital | 83 | 65 (78) | 18 (22) |  |
| Other | 71 | 52 (73) | 19 (27) |  |
| Government | 31 | 23 (74) | 8 (26) |  |
| Retired | 25 | 22 (88) | 3 (12) |  |
| Emergency Room | 20 | 17 (85) | 3 (15) |  |
| Military | 10 | 5 (50) | 5 (50) |  |
| (1361 total practice settings) |  |  |  |  |
| | | | | $\chi^2 (1) = 3.97, p = .046$ |
| Sample sizes for individual analyses varied slightly due to missing data.<br><sup>a</sup>Respondents selected all that apply. |  |  |  |  |

### Self-rated Knowledge & PRS Result Interpretation

Among CAP who reported awareness of PRS, over three-quarters (77%) rated their knowledge about PRS as poor or very poor, while the remaining 23% rated their knowledge as good or very good. Practice site was related to level of self-reported knowledge, with almost one-third (32.6%) of CAP in UMC settings reporting good or very good knowledge of PRS, as compared to only 20% of respondents in non-UMC settings (OR=1.93, 95% CI=1.32 – 2.82, p<.001). There was no significant association (OR=.78, 95% CI=.54 – 1.13, p=.186) between CAP in private practice settings who reported good to very good knowledge of PRS (27%) as compared to CAP who were not in a private practice setting (24.5%) Finally, ratings of good or very good knowledge were not related to length of time in practice (OR=.88, 95% CI=.60 – 1.29, p=.522) when comparing persons with <11 years of practice (24.6%) to persons with ≥11 years of experience (22.3%). Only 26.8% (n=177) of participants correctly answered all five questions when interpreting the example PRS result graph from impute.me. The average number of items correct among all CAP was 3.45 (*SD*=1.57), and the average number of items correct among those who got one or more question wrong was 2.88 (*SD*=1.47).

### Experience

Despite their reported poor knowledge about PRS, some respondents are beginning to see PRS in their clinical practice, with 10.1% (n=74) indicating that they had a patient or a family bring them PRS that they had obtained without the clinician’s involvement, and 4.1% reported they recommended testing for psychiatric PRS in the previous 12 months. Additionally, 4.1% of respondents reported that they had requested or generated psychiatric PRS for a patient. When asked how those PRS were generated, 40% reported they were obtained from a research study, 28.8% from a direct-to-consumer provider, 11.4% from an online tool (e.g., impute.me), and 14.3% indicated they did not know how the PRS were generated. Seven percent of respondents also reported that a patient or their parent or guardian had asked them about psychiatric PRS in the previous 12 months.

### Perceived Utility & Concerns

More than half of respondents thought that PRS have some utility in child and adolescent psychiatry, with 54.1% indicating they thought PRS were *currently* at least slightly useful (see Figure 2). They also believed that PRS will become more useful in the future, with significantly more respondents (86.7%) noting that psychiatric PRS will be at least slightly useful in child and adolescent psychiatry *in five years* (X2 (1) = 220.16, p<.001). When compared to how useful they thought other genetic testing was, they rated the current utility of psychiatric PRS far lower than genetic testing for autism spectrum disorder (ASD; X2 (1) = 182.72, p<.001), for intellectual disability (IDD; X2 (1) = 243.37, p<.001), and for psychiatric conditions other than ASD/IDD (X2 (1) = 107.58, p<.001).

**Fig 2.**
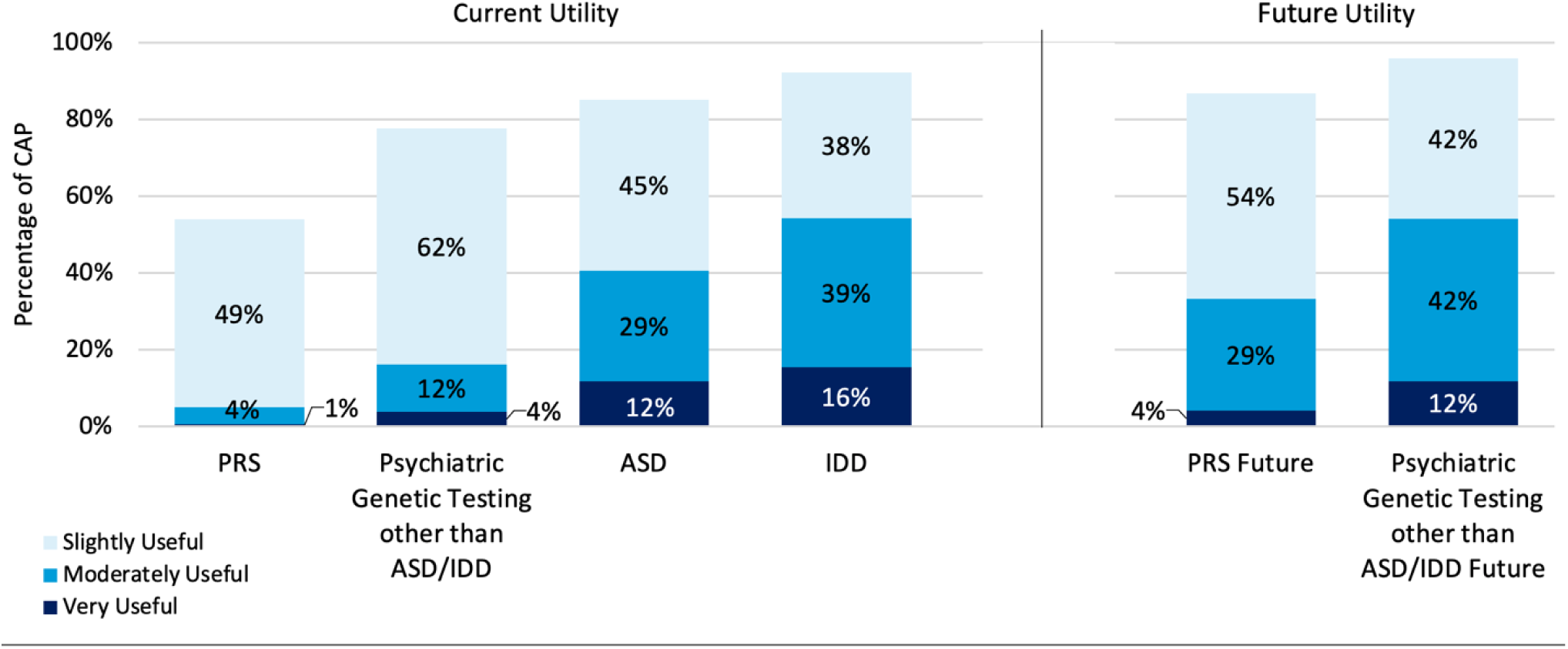
Percentage of CAP who Endorsed any Level of Utility for Each Type of Psychiatric Genetic Testing. Respondents rated the utilty of each type genetic testing on a 4-point scale: Not at all Useful, Slightly Useful, Moderately Useful, Very Useful, PRS=Polygenic Risk Score: IDD=Intellectual/Development Disabilty; ASD=Autism Spectrum Disorder; Future= “in five years”

Respondents, however, also endorsed several concerns that PRS could lead to negative outcomes (Figure 3). A strong majority of respondents indicated they were at least slightly concerned about all listed potential negative outcomes (>89% of respondents for each).

**Fig. 3.**
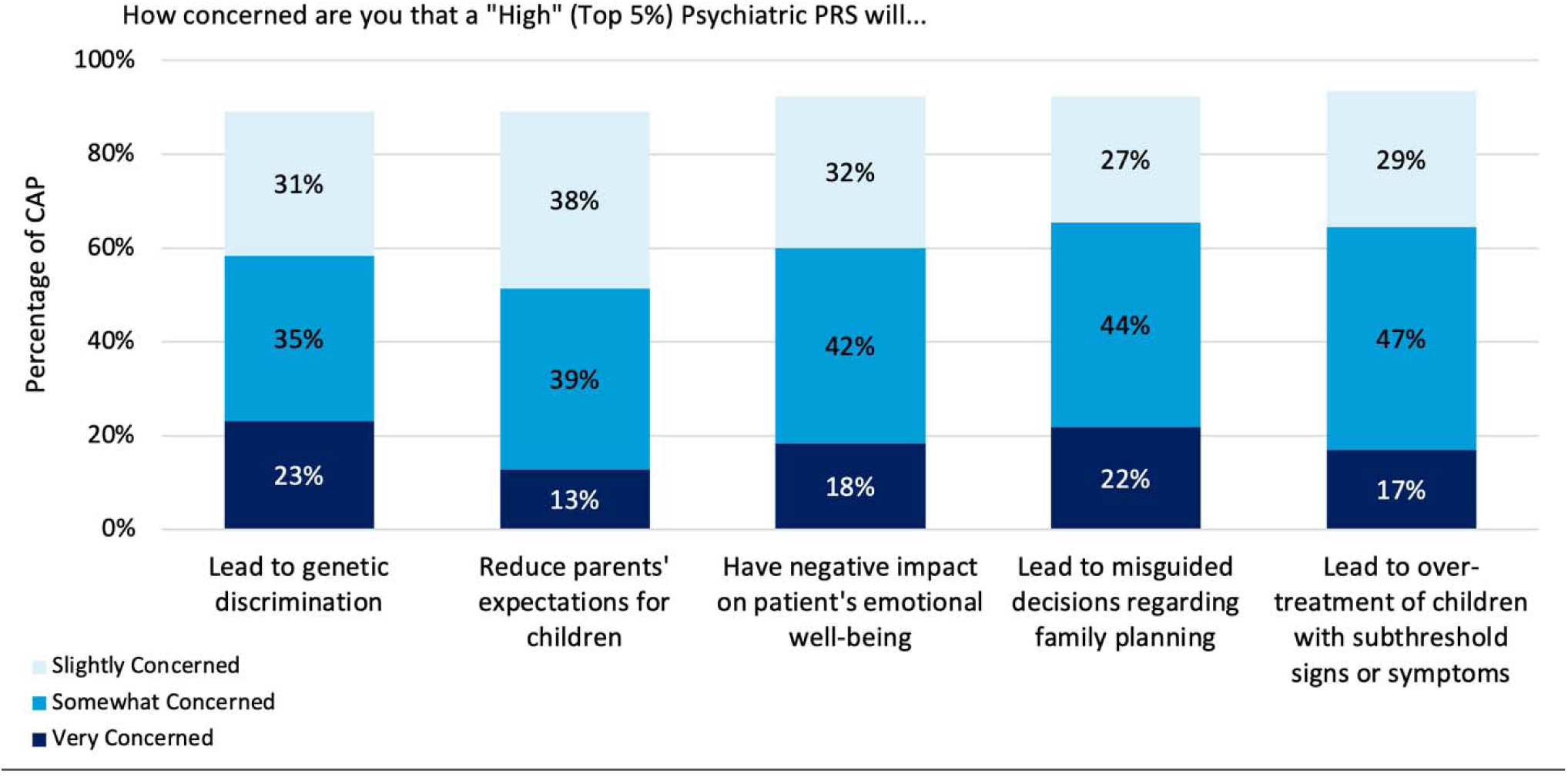
Percentage of CAP Who Endorsed any Level of Concern that a “High” (Top 5%) Psychiatric PRS Could Cause Each Outcome. Respondents rated their level of concern on a 4-point scale: Not at all Concerned, Slightly Concerned, Somewhat Concerned, Very Concerned

### CAP Potential Use of PRS

A third of respondents (34.6%) indicated that nothing would prompt them to request or generate a patient’s psychiatric PRS, while a number of participants endorsed several motivations: 24.9% reported that they would request or generate a patient’s psychiatric PRS if requested by a patient or parent/guardian, 20.5% in response to refractory symptoms, 20.6% for diagnostic clarification, 15.1% to assess risk for a psychiatric condition when a patient has signs or symptoms, 13.6% to assess risk for a psychiatric condition when an asymptomatic patient has a family history, and 12.5% for medication side effects.

CAP were asked what they would do in response to receiving a high (top 5^th^ percentile) psychiatric PRS result for a child or adolescent *with no current psychiatric diagnosis*. Almost half of respondents (47.5%) indicated they would increase monitoring of symptoms, 42.1% would evaluate the patient for symptoms related to psychiatric disorders, 32% would use online resources to learn more, 17.2% would request a consult from a genetic specialist (e.g., genetic counselor, medical geneticist), 17.6% would recommend modifying aspects of the child or adolescent’s life to decrease stress, and 3.9% would prescribe medications to help decrease risk. We also examined the differences in these endorsed actions between those who self-rated their knowledge about PRS as good or very good vs. poor or very poor, and between those who answered all items on the PRS graph interpretation measure correctly vs. those who answered one or more questions incorrectly (Figure 4). Respondents with self-reported good to very good knowledge about PRS were more likely to report they would prescribe medication to reduce risk (OR=2.46, 95% CI=1.16 – 5.28, p=.019) or would do nothing (OR=2.84, 95% CI=1.73 – 4.66, p<.001). Respondents with self-reported good to very good knowledge about PRS were less likely to report they would use online resources to learn more (OR=.41, 95% CI=.27 – .63, p<.001). Respondents who answered all items correctly on the PRS graph interpretation measure were more likely to report that they would evaluate the patient for symptoms related to psychiatric disorders (OR=1.55, 95% CI =1.10 – 2.18, p =.011) and use online resources to learn more (OR=1.77, 95% CI=1.24 – 2.51, p=.002), and were less likely to report they would prescribe medications to help decrease risk (OR=0.23, 95% CI =0.05 – 0.96, p =.044).

**Fig. 4.**
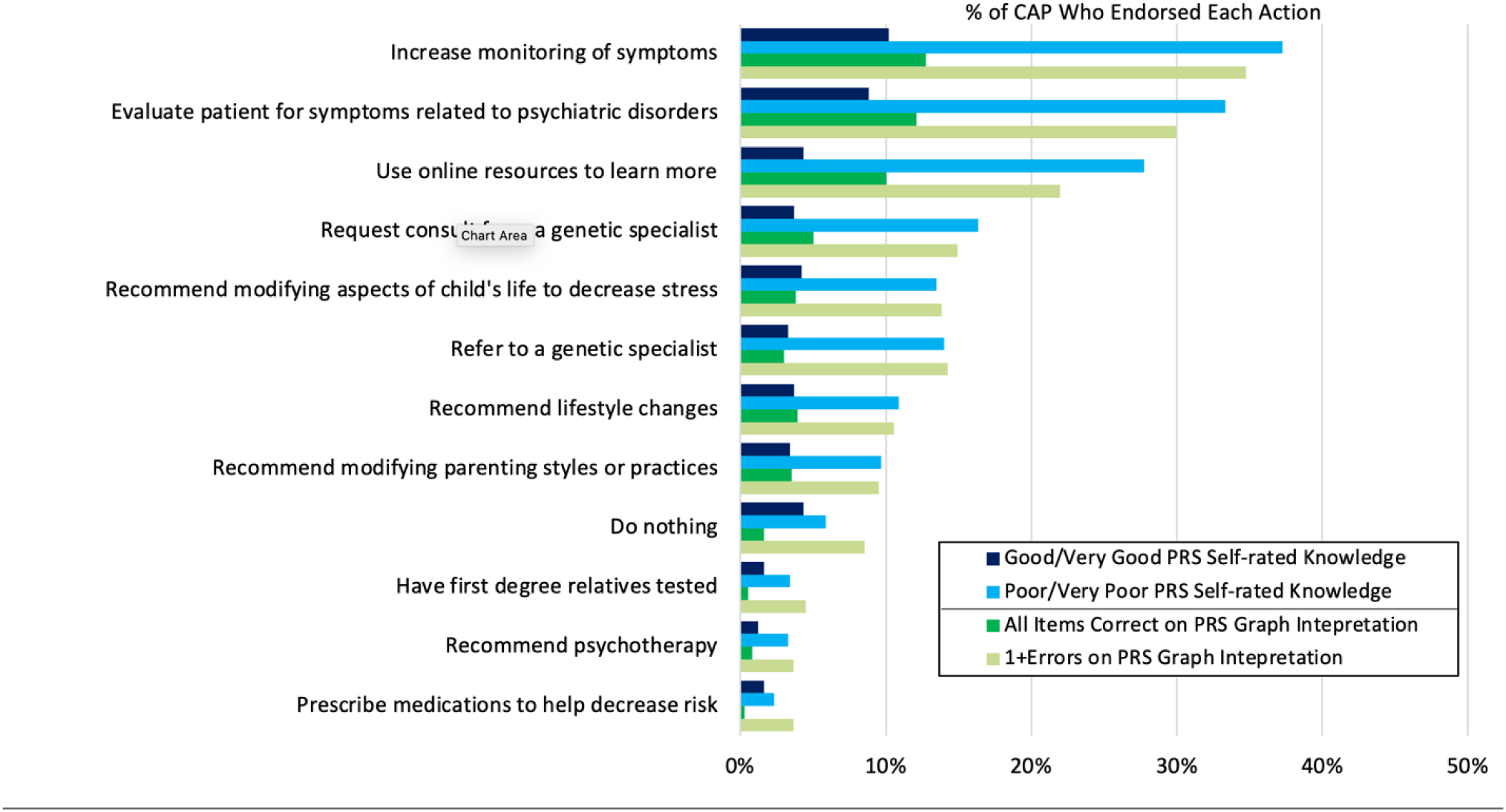
CAP Endorsed Hypothetical Actions in Response to a High (Top 5%) Psychiatric PRS in a Child or Adolescent with No Current Psychiatric Diagnosis by Self-Rated Knowledge and PRS Graph Interpretation.

### CAP Attitudes toward PRS Testing and Psychiatric PRS Screening

We assessed CAP’s attitudes toward using PRS testing in a range of patients and family members for whom psychiatric PRS could potentially be used in the future. About half of respondents felt that it was appropriate or very appropriate to screen first degree relatives of patients diagnosed with a psychiatric disorder *and* a high (top 5^th^ percentile) psychiatric PRS (58.6% of respondents), and first episode patients (48.3%). More than a third felt it was appropriate or very appropriate to screen children or adolescents with sub-threshold signs or symptoms of a disorder (34.6%) and first-degree relatives of an asymptomatic patient with a high PRS (24.8%). There were no group differences in responses about the appropriateness of PRS testing between those who self-rated their knowledge of PRS as good or very good vs. poor or very poor.

Finally, CAP were largely unsupportive of using PRS testing to screen for risk of psychiatric conditions in other contexts, with 90.3% of respondents indicating it was inappropriate or very inappropriate to screen embryos for psychiatric PRS, and 95.7% indicating it was inappropriate or very inappropriate to screen the general population for psychiatric PRS.

## Discussion

In this study of child and adolescent psychiatrists’ knowledge, experience, and attitudes toward the use of PRS, we found that some respondents have already encountered PRS in their clinical practice. Further, most respondents thought that PRS already have some utility and anticipated PRS will have greater utility in child and adolescent psychiatry in the future. Many also reported that a number of scenarios would prompt them to generate a patient’s psychiatric PRS, and many would take actions in response to an undiagnosed child or adolescent’s high psychiatric PRS. While respondents also endorsed multiple concerns and a general low level of knowledge of PRS, these results suggest that CAP may be open to clinical implementation of psychiatric PRS.

Though few respondents have already generated, requested, or recommended testing for psychiatric PRS for a patient, some CAP reported that patients and families are beginning to ask about PRS. With increasing interest around PRS in both academic and popular literature,^33,34^ it is likely CAP will soon see more patients and families asking about PRS. Given that a quarter of our sample reported they would request or generate a patient’s psychiatric PRS if asked to do so by a patient or their parent/guardian, some CAP may be implementing psychiatric PRS into their clinical practice in the not-too-distant future. Further, even if they do not generate PRS or recommend the testing themselves, it is now relatively simple to obtain PRS online using raw data from direct-to-consumer genetic testing (i.e., by uploading raw data to websites like impute.me).^30,31^ Accordingly, some respondents reported that they have had patients or their families bring PRS to them, some of which they reported were generated online.

Respondents also indicated that they would take a number of actions in response to a high PRS for a hypothetical child or adolescent who did not yet have a psychiatric diagnosis, yet the current reliability and predictive power of psychiatric PRS are modest and vary between disorders. Some of the actions endorsed by respondents may be reasonable, such as increasing monitoring of symptoms, evaluating the child for symptoms related to psychiatric disorders, or using online resources to learn more. Other actions, however, may raise concerns, including recommending modifying the child’s life to decrease stress or modifying parent styles or practices, or prescribing medications to help decrease risk. Interestingly, the top concern among respondents was the risk of high PRS leading to over-treatment of children with subthreshold signs or symptoms. However, those who reported greater self-rated knowledge about PRS, and thus may feel more confident making clinical decisions based on PRS, were more likely to indicate they would recommend medications to decrease risk for children with no diagnosis but a high psychiatric PRS.

Given respondents’ overall self-reported low level of knowledge of PRS and poor performance interpreting an example PRS result graph alongside their endorsements of several actions that may be less appropriate, it is imperative that further research explore how these PRS are currently being used and best practices for implementation into child and adolescent psychiatry care. In particular, these data highlight the largely untapped opportunity for interdisciplinary collaboration between psychiatrists and clinical genetics professionals in this area^35^ (genetic counselors in different geographical areas with various areas of expertise can be found using the www.findageneticcounselor.com tool). Importantly, genetic information has the potential for positive or negative reactions from patients and their families. Indeed, the manner in which this information is delivered is crucial in determining outcome.^36^ A collaborative approach between psychiatry and clinical genetics would ensure the implementation of any genetic testing information in the context of evidence based psychiatric genetic counseling, which has been shown to result in positive patient outcomes.^14^

Given that respondents did not rate the current utility of psychiatric PRS very high, it was surprising to see the level of support for PRS testing across a range of groups, including first episode patients and children or adolescents with subthreshold symptoms. Respondents, however, were far less supportive of using psychiatric PRS testing to screen the general population, or to screen embryos generated via *in vitro* fertilization cycles. As noted elsewhere,^37^ polygenic embryo screening amplifies a number of ethical and policy issues already present in screening embryos for monogenic conditions, and it also poses new challenges, including the ability to screen for risk for psychiatric conditions, as well as traits (e.g., height, intelligence).^38–40^

Our results should be interpreted within several limitations. Though we successfully surveyed 11.6% of CAP in the US, our sample may not be representative of the larger group, and it is possible there may be response bias in our sample. For instance, those who had never heard of PRS may have been less likely to respond, or those with more favorable views toward PRS may have been more likely to participate. We did not collect data from those who did not respond to the survey, thus we are unable to assess whether respondents differed meaningfully from non-responders. Some questions, such as the self-rated knowledge question, may be prone to social desirability bias. Despite these limitations, this survey provides critical insight into CAP knowledge, experience, and attitudes toward the use of psychiatric PRS in child and adolescent psychiatry, and as such, it can inform needs for future research and best practices.

Given the rapidly emerging use of psychiatric PRS in different contexts, including child and adolescent psychiatry, and the growing numbers of samples of psychiatric GWAS, which will give rise to more reliable PRS for more psychiatric disorders, it is critical to begin educational efforts for clinicians and patients to examine the ethical and clinical challenges that PRS may generate, and for relevant professional organizations to develop practice recommendations to help guide clinicians as to how to use these tools. Our findings highlight some of the most pressing issues and underscore the urgency to take action.

## Data Availability

All data produced in the present study are available upon reasonable request to the authors.

## Data Availability

De-identified survey data are available on request from the corresponding author. Data will be made available to researchers whose proposed use of the data has been approved by the study team.

## Ethics Declaration

The BCM Institutional Review Board (IRB) approved this study (protocol number: H-46219). Informed consent was obtained from all participants as required by the BCM IRB.

## Acknowledgments

This work was funded by the National Human Genome Research Institute and the National Institute of Mental Health of the National Institutes of Health, grants R00HG008689 (Lázaro-Muñoz), 3R00HG008689-05S1 (Lázaro-Muñoz, Storch), R01MH128676 (Pereira, Lázaro-Muñoz, Storch), and P50HD103555 (Storch) for use of the Clinical and Translational Core facilities. The views expressed are those of the authors alone, and do not necessarily reflect views of NIH, Baylor College of Medicine, or Harvard Medical School.

